# Same Result, Different Price: Compounded versus Branded Tirzepatide

**DOI:** 10.64898/2026.07.14.26357505

**Authors:** Brian Erly, Shanmugesh Raja

## Abstract

**Background:** Compounded tirzepatide is prescribed at scale as a cheaper substitute for branded Mounjaro and Zepbound, yet the cost case is almost always built by setting one compounded price against one branded list price. That framing ignores the question that actually decides the answer: cheaper than which branded price the patient can reach. Branded tirzepatide is now sold at sharply different tiers, namely insurance copay (often $25–$150/month), LillyDirect Self Pay ($299–$449/month), and retail cash price ($1,000–$1,200/month). Whether compounded saves money turns entirely on which of these a given patient faces. A second open question is whether the two formulations even produce comparable effectiveness, since observed differences may reflect selection on insurance, baseline characteristics, and adherence rather than the drug.

**Methods:** We conducted a retrospective cohort study of tirzepatide users in the Mochi Health tele-health program, classified by formulation from their refills as branded-only (Mounjaro/Zepbound; 6,238), compounded-only (71,683), or switchers (4,996); switchers were excluded from the formulation contrast. Among single-formulation patients with a documented six-month weight observation, the analytic cohort was 7,271 (869 branded, 6,402 compounded). The primary outcome was six-month percent body weight loss; the secondary outcome was ≥ 10% response. We used 1:1 nearest-neighbor propensity-score matching (0.25 SD caliper) on baseline covariates only — age, sex, baseline BMI, baseline weight, comorbid diabetes, hypertension, dyslipidemia, prior bariatric surgery, and self-reported insurance coverage — deliberately excluding post-treatment variables such as adherence and time in program, which are mediators of the formulation effect. We pre-specified an equivalence margin of ± 2 percentage points on mean loss and tested equivalence with two one-sided tests (TOST). A directed acyclic graph (DAG) makes the identifying assumptions explicit; metformin use could not be reliably ascertained and is treated as an unmeasured confounder. The cost comparison reports the savings or premium of compounded versus branded under five branded price scenarios: retail list, LillyDirect Self Pay (two dose tiers), and insurance copay (typical and low end). It is a cost comparison (cost-minimization under demonstrated similar effectiveness), not a formal cost-effectiveness analysis: we computed no ICER, QALY, or discounting.

**Results:** Branded and compounded patients had similar outcomes even before adjustment (mean loss 11.7% vs 11.5%; ≥ 10% response 60.9% vs 58.8%). The largest baseline difference between the groups was insurance coverage (branded patients far more likely insured; standardized mean difference 0.67), which matching balanced to 0.01. After 1:1 matching (718 pairs, all |SMD| < 0.04), mean loss was 11.4% vs 11.4% (difference +0.08 pp, 95% CI −0.70 to +0.80) and ≥ 10% response 59.3% vs 57.2% (difference +2.1 pp, 95% CI −3.1 to +7.1). The two formulations were statistically equivalent within the pre-specified ± 2 pp margin (TOST *p* < 0.001). Cost depends on the branded scenario: compounded saves $6,000 over six months versus retail list price, $1,494 versus LillyDirect maintenance-dose (5–15 mg) Self Pay, and $594 over a low-dose (2.5 mg) LillyDirect prescription, while it costs $300 more than branded under a typical insurance copay ($150/month) and is more expensive still at lower copays (savings turn negative below $200/month).

**Conclusions:** Branded and compounded tirzepatide were statistically equivalent in six-month effectiveness within a pre-specified ± 2 pp margin, so the choice between them is essentially a cost decision — and that cost advantage is real but conditional on the branded price the patient can access. It is large against retail list price and shrinks to zero or reverses against LillyDirect Self Pay or a low insurance copay. Whether compounded is the lower-cost choice for an individual patient is, therefore, a question about which price tier that patient faces.

## 1. Introduction

The case for compounded tirzepatide is almost always made on price. A list-price branded prescription runs $1,000 to $1,200 per month; compounded preparations are routinely available for $200 to $400. Stated that way, the savings look decisive. But the comparison answers the wrong question. The economically meaningful question is not whether compounded is cheaper than branded in the abstract, it is whether compounded is cheaper than the branded price the patient can actually reach. Branded tirzepatide does not have one price. It has a distribution of prices spanning nearly two orders of magnitude, and the savings claim either holds or collapses depending on which tier the patient occupies. A patient paying $1,200 cash and a patient paying a $25 copay are looking at entirely different decisions, and a single-anchor cost comparison silently assumes the former for everyone.

This paper makes two contributions. First, it tests whether compounded and branded produce comparable effectiveness at all. Unadjusted real-world comparisons consistently show branded out-performing compounded, but that gap is exactly what selection would produce: branded uptake correlates with insurance, with adherence support, and with baseline weight, so raw means confound formulation with the patients who receive it. We use propensity-score matching, supported by an explicit DAG, to separate the drug from the patient. Second, the paper reframes the cost question itself. Rather than report a single savings figure, we estimate the cost premium of branded over compounded across the five price tiers a U.S. patient may actually face in 2026, treating the savings as a function of access rather than a constant.

Both questions have force because the pricing landscape has shifted. Lilly’s direct-to-consumer pharmacy (LillyDirect Self-Pay Journey program, launched February 2026) sells single-dose vials of tirzepatide 2.5 mg at $299/month, 5 mg at $399/month, and 7.5–15 mg maintenance doses at $449/month, well below the retail cash price.^1^ Many insured patients face copays below $200/month. The branded-compounded differential is no longer a number; it is a distribution, and the cost conclusion is conditional on where in that distribution the patient sits.

## 2. Methods

### 2.1 Cohort

This is a retrospective cohort study using electronic health record and pharmacy data from Mochi Health, a U.S. telehealth obesity program. We identified tirzepatide users and classified each by the formulation on their refills as branded-only (Mounjaro/Zepbound; *n* = 6,238), compounded-only (*n* = 71,683), or switchers (*n* = 4,996); switchers were excluded from the head-to-head comparison to avoid confounding the formulation contrast with switching. Among single-formulation patients with a measured weight in the six-month follow-up window (135–225 days after the first refill), the analytic cohort was *n* = 7,271: 869 branded-only and 6,402 compounded-only patients (718 matched pairs after PSM). Baseline weight was the measured weight closest to the first refill within ± 30 days; the six-month weight was the measurement closest to 180 days within the follow-up window.

### 2.2 Outcomes

The primary outcome was mean percent body weight loss at 6 months. The secondary outcome was response rate, defined as ≥ 10% loss at 6 months. FDA guidance on weight-management drug development frames efficacy in terms of categorical responder thresholds (the proportion of patients achieving at least 5% weight loss),^20^ and the ≥10% threshold used here is consistent with the responder benchmarks reported in the pivotal GLP-1 obesity-pharmacotherapy trials.^3,6,8^

### 2.3 Causal-inference framework and DAG

We used propensity-score matching^4^ to address selection on observed covariates and built a DAG to make the identifying assumptions explicit. The DAG (Figure 1) was developed with a clinical co-author (BE) and is consistent with the dagitty implementation.^5^

**Figure 1:**
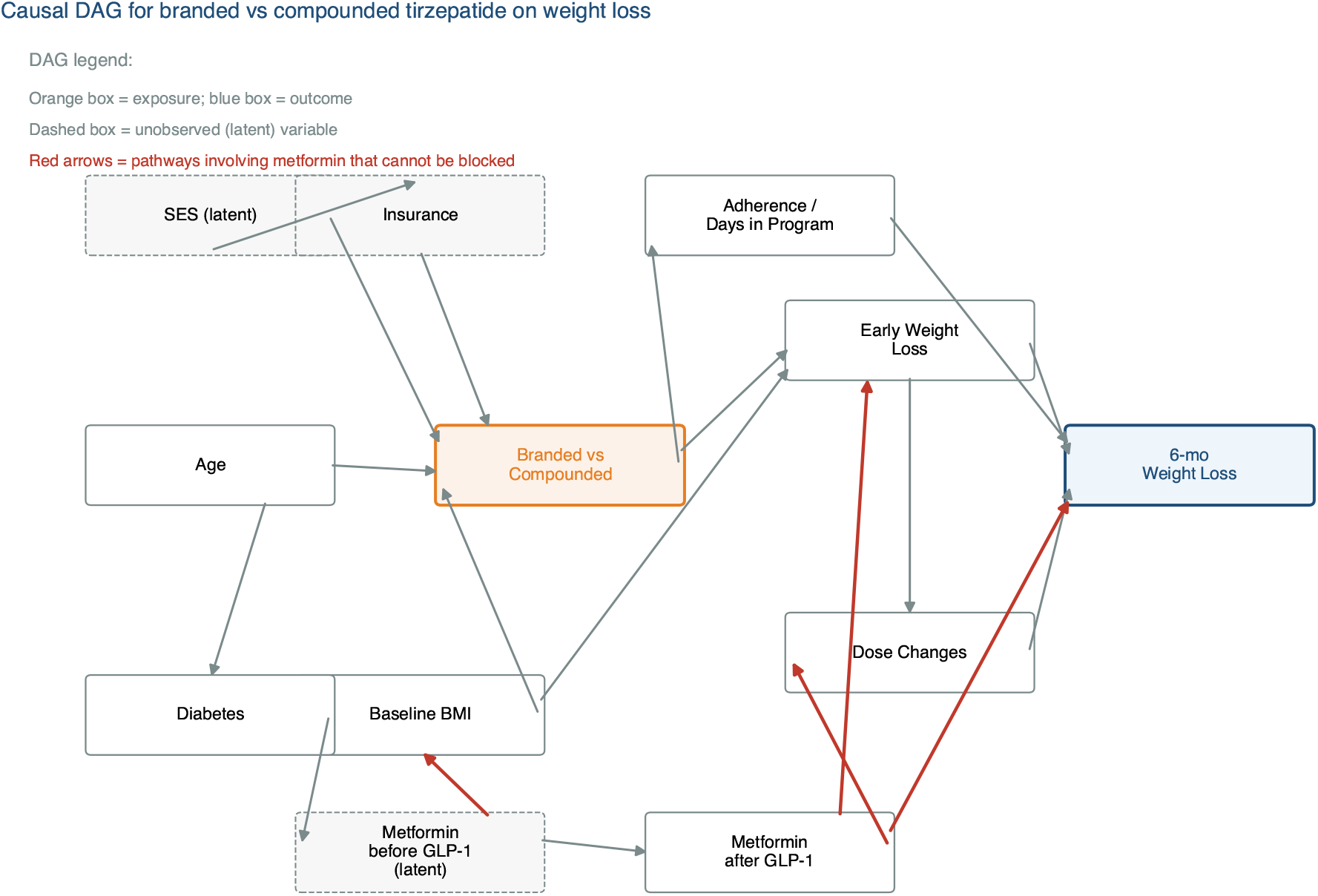
Causal DAG for branded vs compounded tirzepatide on 6-month weight loss. Orange box is the exposure (formulation); blue box is the outcome. White boxes are observed covariates we adjust for; dashed boxes are latent (unobserved). Red arrows mark the metformin-mediated pathways that cannot be blocked: pre-GLP-1 metformin influences both insurance access and baseline BMI (confounder structure), while post-GLP-1 metformin acts simultaneously as a confounder (some patients take it for diabetes) and a mediator (some patients take it as a weight-loss adjunct). Adjusting for “metformin use” without separating these uses introduces bias of unpredictable direction.

The DAG sorts variables into three groups. The **adjusted variables** (age, sex, baseline BMI, baseline weight, comorbid diabetes, hypertension, dyslipidemia, prior bariatric surgery, and self-reported insurance coverage) are observed and enter the propensity model. **Mediators** (adherence, time in program, dose changes, early weight loss) lie on the path from formulation to outcome and are deliberately *not* adjusted, to avoid over-adjustment bias. The **unobserved variables** (true socioeconomic status and concomitant medications such as metformin) cannot be measured in our data; metformin in particular is a confounder when taken for diabetes and a mediator when taken as a weight-loss adjunct, and our data cannot tell the two uses apart, so it is treated as residual confounding (addressed under *Residual confounding* in the Results) rather than adjusted.

### 2.4 Propensity-score matching

We performed 1:1 nearest-neighbor matching without replacement, with a caliper of 0.25 SD on the logit propensity. The propensity model is a logistic regression of formulation (branded = 1, compounded = 0) on baseline covariates only: age, sex, baseline BMI, baseline weight, comorbid diabetes, hypertension, dyslipidemia, prior bariatric surgery, and self-reported insurance coverage. We deliberately excluded post-treatment variables (refill counts, time in program, dose changes), which sit on the causal path from formulation to outcome; adjusting for them would be over-adjustment that biases the formulation contrast. Balance was assessed by standardized mean differences (SMD), treating |SMD| < 0.10 as adequate; matching yielded *n* = 718 branded–compounded pairs. Because equivalence, not difference, is the substantive question, we pre-specified an equivalence margin of ± 2 percentage points on mean six-month loss and tested it with two one-sided tests (TOST); equivalence is declared when both one-sided tests reject at *p* < 0.05 (the 90% CI for the difference lies entirely within the margin).

### 2.5 Insurance as a matched covariate

Insurance is the covariate most likely to drive selection between formulations: insured patients can often reach branded at a copay, whereas uninsured patients more often turn to compounded. We therefore included a self-reported insurance-coverage indicator directly in the propensity model rather than handling it post hoc, and we report its balance before and after matching (Table 1).

**Table 1:** Covariate balance before and after matching. (standardized mean differences). Insurance, the dominant imbalance, is balanced after matching.

| Covariate | SMD before | SMD after |
| --- | --- | --- |
| Age | −0.49 | 0.02 |
| Female sex | −0.10 | 0.02 |
| Baseline BMI | 0.00 | −0.01 |
| Baseline weight | 0.00 | −0.02 |
| Comorbid diabetes | 0.05 | −0.01 |
| Hypertension | −0.08 | −0.01 |
| Dyslipidemia | −0.12 | −0.01 |
| Prior bariatric surgery | −0.08 | 0.04 |
| Self-reported insurance | 0.67 | 0.01 |

**Table 2:** Effectiveness comparison (mean six-month percent loss). The matched difference (+0.08 pp, 95% CI −0.70 to +0.80) is statistically equivalent within the pre-specified ±2 pp margin (TOST *p* < 0.001).

| Analysis | Branded loss (%) | Compounded loss (%) | $\Delta$ (pp) [95% CI] |
| --- | --- | --- | --- |
| Unadjusted (869 vs 6,402) | 11.71 | 11.51 | +0.20 |
| PSM-matched (718 pairs) | 11.44 | 11.37 | +0.08 [−0.70, +0.80] |

### 2.6 Cost analysis: price scenarios

The standard cost comparison anchors branded at retail list price (approximately $1,200/month for a typical 5–10 mg dose) and compounded at the program’s median price (approximately $200/month). This is a cost comparison under the assumption of similar effectiveness (a cost-minimization framing), not a formal cost-effectiveness analysis; we compute no ICER, QALY, or discounting. We extend this to five branded scenarios spanning the price tiers a 2026 U.S. patient may actually face:

1. Retail list price (∼$1,200/month)
2. LillyDirect Self Pay 7.5–15 mg maintenance vial ($449/month)^1^
3. LillyDirect Self Pay 2.5 mg vial ($299/month)^1^
4. Branded with insurance copay, typical ($150/month)
5. Branded with insurance copay, low end ($25/month)

For each scenario we compute the 6-month cost premium of branded over Mochi compounded ($200/month × 6 = $1,200) and report the savings of compounded conditional on the comparable effectiveness suggested by PSM. All savings are computed against this single $200/month compounded anchor; if a higher compounded price were used (for example the cited market-median of approximately $300/month) every savings figure would shrink correspondingly and the negative-savings (“no case”) regime would widen. To be plain about the choice of anchor: compounded tirzepatide trades across an approximately $200–$400/month range, with a market median near $300/month, and the savings reported here use the $200/month (low end) anchor rather than the median; readers should note that the savings figures would be smaller at the $300 median and smaller still toward the top of the range, so these estimates are not built at the least favorable compounded price.

### 2.7 What we did not do, and why

We did not run a formulation-additive subanalysis comparing compounded preparations with B12, glycine, niacinamide, or pyridoxine additives. The additive doses in compounded tirzepatide are sub-therapeutic with short half-lives, and no biological mechanism supports an effect on weight loss at those concentrations. In our data, “additive type” most plausibly fingerprints the compounding pharmacy rather than a pharmacologically meaningful exposure, and a credible by-pharmacy comparison would require pharmacy-level identifiers we do not have. We therefore drop the subanalysis.

### 2.8 Software

Python 3.12 with scikit-learn 1.9, pandas 2.3, statsmodels 0.14, and SciPy 1.17. Matched-difference confidence intervals were obtained by 2,000-replicate bootstrap; equivalence was assessed by two one-sided tests (TOST) against a pre-specified *±*2 pp margin.

## 3. Results

### 3.1 Baseline characteristics and balance

The branded and compounded groups were similar on most baseline characteristics; the one large difference was insurance coverage, with branded patients far more likely to report it (standardized mean difference 0.67). After 1:1 PSM on the nine baseline covariates, every standardized mean difference fell below 0.04 in the matched sample of 718 pairs (Table 1), insurance included (0.67 before matching, 0.01 after).

### 3.2 Effectiveness: unadjusted, PSM-matched, and equivalence

Branded and compounded produced similar outcomes both before and after adjustment (Figure 2). Unadjusted, branded led by only +0.20 pp on mean loss (11.71% vs 11.51%) and +2.1 pp on response (60.9% vs 58.8%). After matching, mean loss was 11.44% vs 11.37% (difference +0.08 pp, 95% CI −0.70 to +0.80) and ≥ 10% response was 59.3% vs 57.2% (difference +2.1 pp, 95% CI −3.1 to +7.1); neither difference was statistically significant. Critically, the matched mean-loss difference satisfied the pre-specified equivalence test: both one-sided tests rejected at *p* < 0.001, so branded and compounded are statistically *equivalent* within the ± 2 pp margin, not merely “not significantly different.” The small unadjusted edge reflects who chooses branded, chiefly the more-insured patients, rather than the formulation.

**Figure 2:**
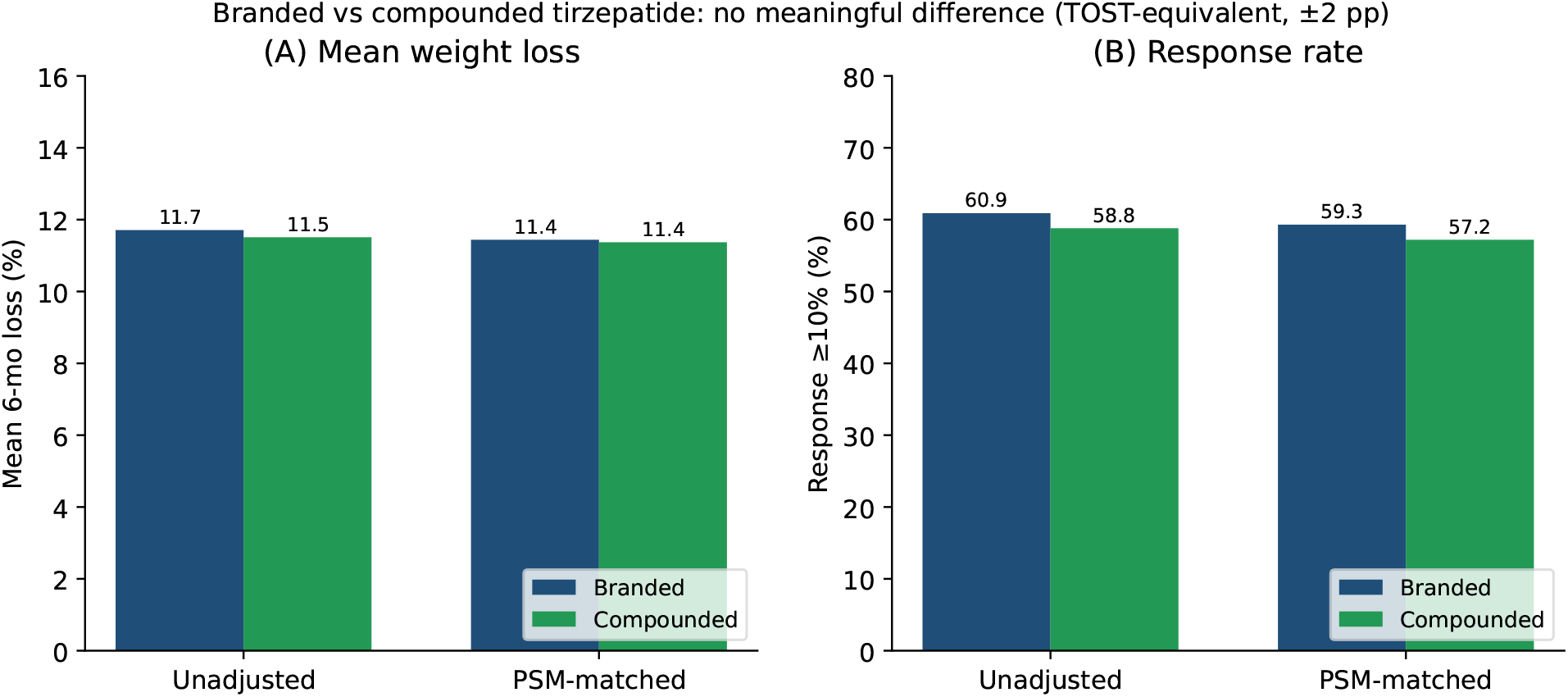
Effectiveness comparison, unadjusted vs PSM-matched. (A) Mean six-month percent body weight loss. (B) Response rate (≥ 10% loss). Branded and compounded are similar in both views; the matched mean-loss difference is statistically equivalent within the pre-specified ± 2 pp margin (TOST *p* < 0.001). The small unadjusted edge reflects selection on insurance, not formulation efficacy.

### 3.3 Cost: no detected outcome difference, savings depend on branded price tier

Once we treat branded and compounded as producing similar 6-month outcomes (no significant difference detected, Section 3.2), the cost comparison reduces to a single question: which formulation costs the patient less. The answer depends entirely on which branded price tier the patient can access (Figure 3, Table 3).

**Table 3:** Six-month cost premium of branded over Mochi compounded ($1,200 / 6 mo).

| Branded scenario | Branded 6-mo cost | Premium over compounded |
| --- | --- | --- |
| Retail list price (\$1,200/month) | \$7,200 | +\$6,000 |
| LillyDirect 7.5–15 mg (\$449/month) | \$2,694 | +\$1,494 |
| LillyDirect 2.5 mg (\$299/month) | \$1,794 | +\$594 |
| Branded copay typical (\$150/month) | \$900 | –\$300 |
| Branded copay low end (\$25/month) | \$150 | –\$1,050 |

**Figure 3:**
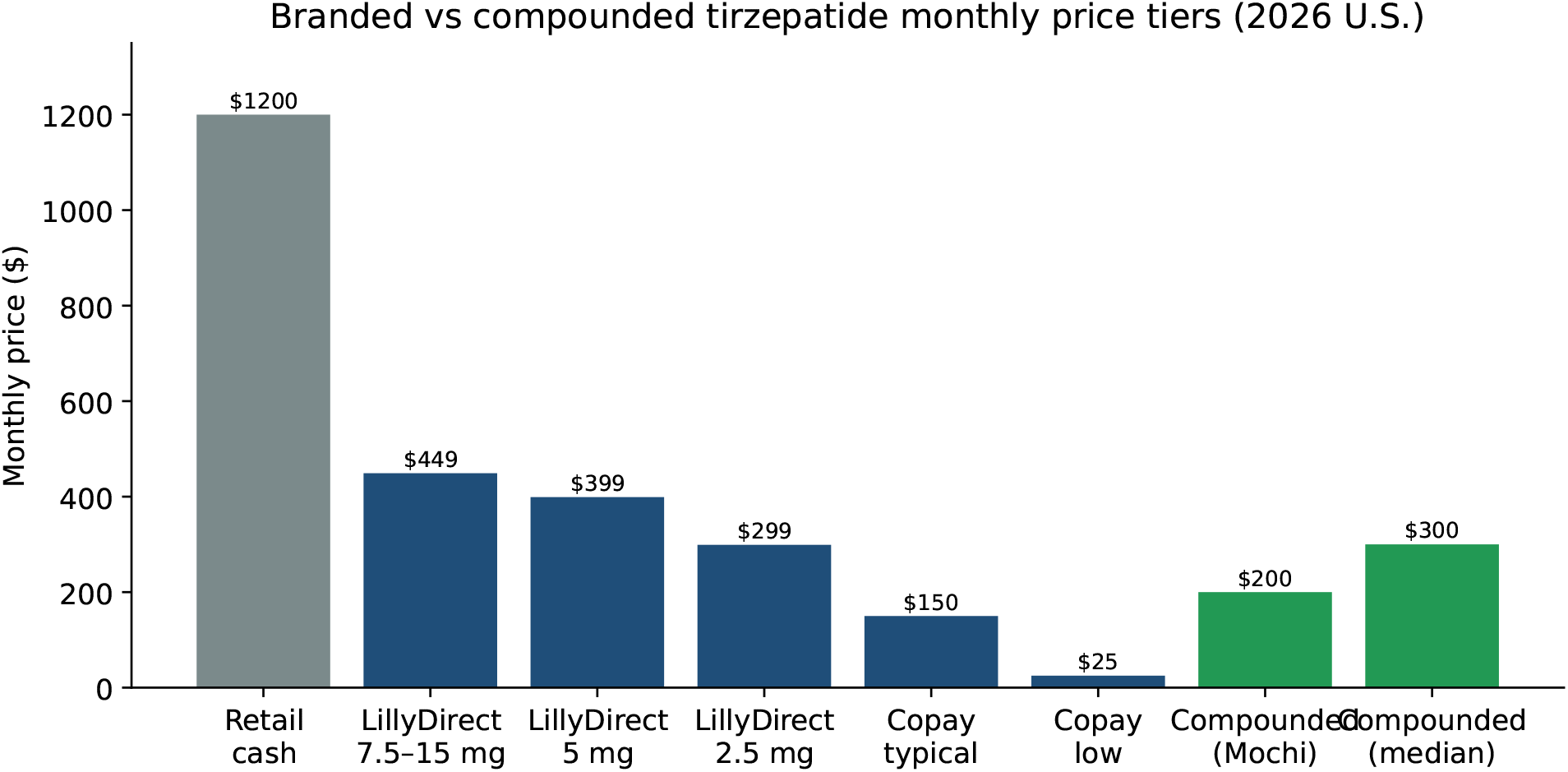
Branded and compounded tirzepatide price tiers (2026 U.S.). Branded prices range across nearly two orders of magnitude depending on the patient’s coverage and access pathway. Mochi compounded sits at $200/month; market-median compounded prices are around $300/month.

**Figure 4:**
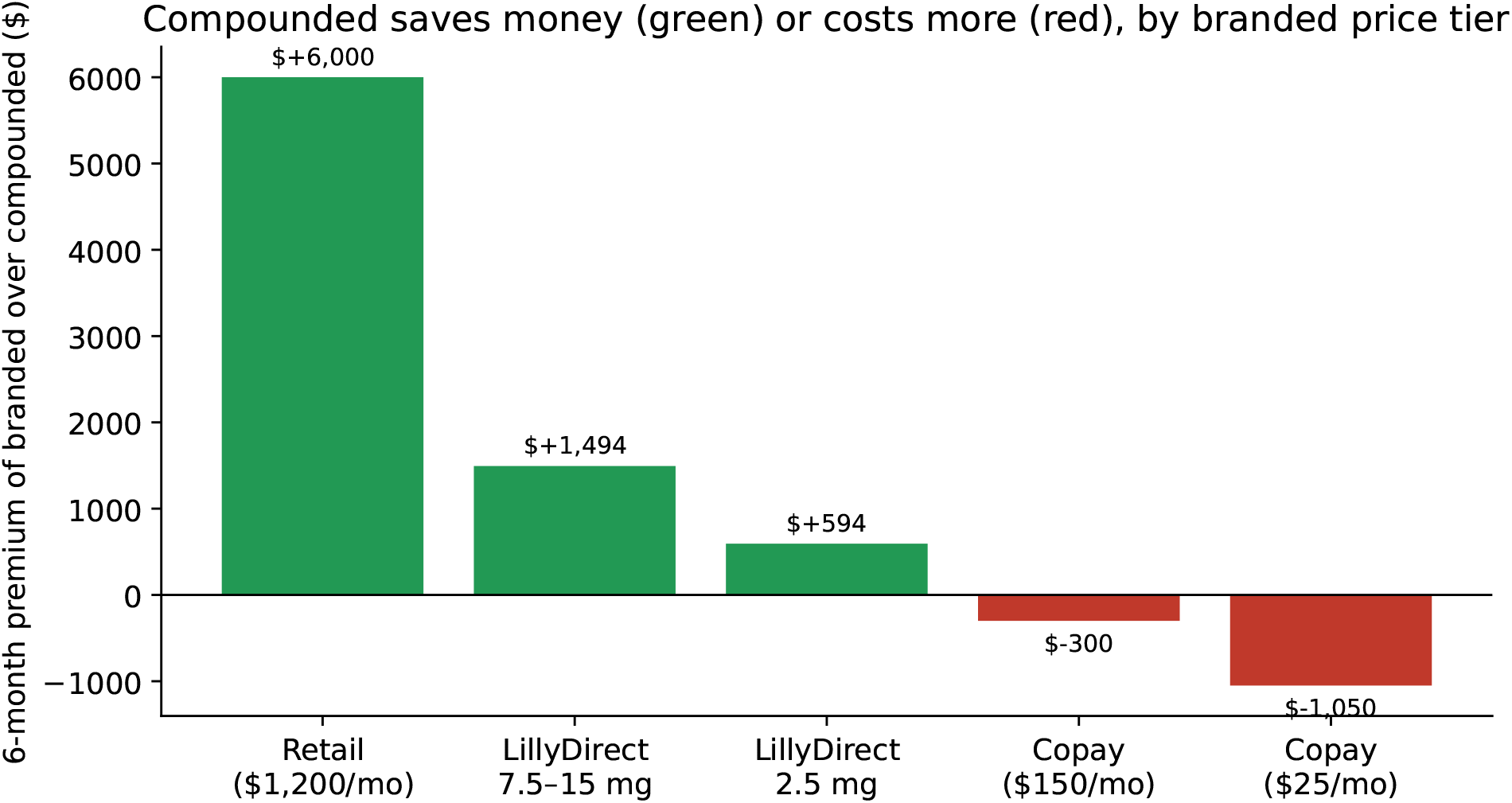
Six-month cost premium of branded over Mochi compounded. Green bars: branded costs more than compounded (compounded saves money). Red bars: branded costs less than compounded (compounded is more expensive). The cost-savings case for compounded shrinks dramatically as the patient’s branded price access improves.

### 3.4 Where compounded saves money, where it does not

The cost story splits into three regimes. In the **strong case**, patients without insurance and without LillyDirect access face the retail list price of approximately $1,200/month, and compounded saves approximately $6,000 over a six-month course. This is the regime in which the standard cost claim is correct. In the **modest case**, patients without insurance who use LillyDirect Self Pay save approximately $594–$1,494 over six months by choosing compounded, depending on dose ($594 at the 2.5 mg tier, $1,494 at the maintenance-dose tier per Table 3); the savings are real but substantially smaller than the retail comparison implies. In the **no case** regime, patients with an insurance copay below $200/month pay more out of pocket for compounded than for branded, so the comparison favors branded and compounded becomes a more-expensive substitute for a product with no detected effectiveness difference.

### 3.5 How many U.S. patients actually face each regime

Because the cost conclusion depends on the regime, the population-level case for compounded depends on how many U.S. adults fall into each one. For 2024–2026, the honest picture is that being well-insured for obesity pharmacotherapy is a minority status, not the default.

#### Uninsured

An estimated 25.3 million U.S. adults were uninsured at any point in 2023 (U.S. Census Bureau).^9^ These patients face retail list price for branded, and compounded is the only way most of them access tirzepatide at scale.

#### Underinsured and high-deductible

The Commonwealth Fund’s 2024 Biennial Health Insurance Survey found that 44% of working-age adults with continuous insurance coverage were underinsured (out-of-pocket costs were 10% or more of income, or the deductible was 5% or more of income).^10^

The same survey found 23% of insured adults had a deductible of $2,000 or more.^10^ A patient with a $2,500 deductible and 30% coinsurance does not face the $25 copay scenario; until the deductible is met they face something close to retail list.

#### Coverage of GLP-1 for obesity specifically

Even among the insured, coverage of branded GLP-1 medications for obesity, as opposed to diabetes, is uneven. As of 2024, KFF reports that fewer than half of large U.S. employers covered semaglutide or tirzepatide for obesity without additional restrictions, and Medicare does not cover any GLP-1 receptor agonist for obesity absent an FDA-approved cardiovascular indication.^11,12^ Patients in plans without obesity coverage face retail list price for branded regardless of their overall insurance status.

The implication is direct: the “no case” regime applies to a relatively narrow subset of patients, namely commercially insured patients with active obesity coverage and low copays. The “strong case” and “modest case” together cover the larger fraction of the U.S. obesity-pharmacotherapy market, and for high-deductible patients compounded is often the only practical pathway to therapeutic-dose GLP-1 access in the first six months of care.

### 3.6 Access to GLP-1 therapy matters beyond weight loss

The access argument changes the comparison itself. Obesity is a chronic, relapsing disease for which clinical guidelines endorse sustained pharmacotherapy, and in which interrupting treatment is associated with weight regain,^2,7^ so for a substantial fraction of patients the realistic choice is not branded versus compounded for weight loss; it is GLP-1 therapy via compounded versus no GLP-1 therapy at all. That shifts the relevant outcome distribution, because GLP-1 receptor agonists carry established benefits beyond weight loss that have their own economic and clinical value.

#### Important caveat on extrapolation

The cardiovascular, heart-failure, and renal trials cited below studied *branded semaglutide* (and, for sleep apnea, branded tirzepatide), not the compounded tirzepatide examined in this paper. They measured cardiovascular, renal, and symptom outcomes that this study did not measure and was not designed to assess. Whether these benefits transfer to compounded tirzepatide is an unproven extrapolation that rests on assumptions about pharmacologic class effect and formulation comparability we cannot test with our data. The discussion below is therefore presented as context for the access argument, not as evidence that compounded tirzepatide confers these specific outcomes.

#### Cardiovascular outcomes

The SELECT trial randomized 17,604 adults with overweight or obesity (no diabetes) and pre-existing cardiovascular disease to semaglutide 2.4 mg/wk or placebo and reported a 20% relative reduction in major adverse cardiovascular events over a mean follow-up of 39.8 months (HR 0.80, 95% CI 0.72–0.90).^13^

#### Heart failure with preserved ejection fraction

The STEP-HFpEF trial randomized 263 adults with HFpEF and obesity to semaglutide 2.4 mg/wk; semaglutide improved Kansas City Cardiomyopathy Questionnaire symptom score by 7.8 points relative to placebo and improved six-minute walk distance.^14^

#### Diabetic kidney disease

The FLOW trial randomized 3,533 patients with type 2 diabetes and chronic kidney disease to semaglutide 1.0 mg/wk or placebo; semaglutide reduced major kidney-disease events by 24% (HR 0.76, 95% CI 0.66–0.88).^15^

#### Obstructive sleep apnea

The SURMOUNT-OSA trials randomized 469 adults with moderate-to-severe OSA and obesity to tirzepatide or placebo and reported substantial reductions in apnea-hypopnea index across both trials.^16^

#### Diabetes prevention

STEP and SURMOUNT secondary analyses have documented reductions in new-onset diabetes in obese patients treated with semaglutide and tirzepatide; in STEP 4 the difference in HbA1c and glycemic indices was substantial.^17^

These outcomes are not the endpoint of this paper, and our data do not measure them; the trials above studied branded products and outcomes outside our scope. The narrower observation is that, to the extent these class benefits apply, “no GLP-1 therapy” may not be a neutral default for a high-deductible patient. We do not claim compounded tirzepatide delivers these specific cardiovascular, renal, or metabolic outcomes, and any such transfer is speculative given the formulation and outcome differences noted above.

### 3.7 Reframing the additive question: tolerability, not weight-loss enhancement

We dropped the formulation-additive subanalysis from the primary comparison (Section 2.7) because the additive doses are sub-therapeutic with short half-lives and no defensible mechanism supports an effect on six-month weight loss at those concentrations. That does not make the option of additive inclusion clinically irrelevant, and three observations are worth recording.

#### Pyridoxine (vitamin B6) has established antiemetic activity

B6 is a long-standing first-line therapy for nausea and vomiting of pregnancy, endorsed by ACOG, and the pyridoxine-doxylamine combination is FDA-approved (Diclegis) for that indication.^18^ Nausea is the most common adverse event leading to GLP-1 discontinuation,^19^ and although pyridoxine has not been tested head-to-head against GLP-1 nausea specifically, its inclusion in a compounded preparation is consistent with the established mechanism for nausea management.

#### B12 (cyanocobalamin) is widely used in weight-loss programs for energy and well-being

Patients on calorie-restricted diets and patients with GLP-1-mediated reductions in food intake may have lower micronutrient intake, and B12 supplementation is a routine adjunct in many obesity-medicine practices. We make no claim that B12 changes weight-loss efficacy in compounded tirzepatide; we note only that the option to include it without a separate prescription is a practical tolerability and adherence feature.

#### Individualized formulation is something branded cannot offer

Compounded preparations can be tailored to the patient: pyridoxine when nausea is a known problem, B12 when intake suppression is a concern, omitting either on preference. Branded tirzepatide is a single SKU per dose tier. The trade-off is regulatory: the FDA-approved branded product is bioequivalent to the trial formulation and carries a more rigorous safety profile, while compounded preparations carry the variability of compounding-pharmacy practice. That variability is real, and it is one reason we did not run a pharmacy-level analysis (Section 2.7).

The honest summary on additives is that we do not claim they enhance weight loss. We do claim that the structural flexibility of compounded prescribing supports tolerability and adherence management the branded product cannot offer, which matters most for patients who would otherwise discontinue the medication.

### 3.8 Residual confounding: metformin and SES

The DAG (Figure 1) flags metformin as a structural confounder PSM cannot fix: it is a confounder when taken for diabetes and a mediator when taken as a weight-loss adjunct, and the two uses cannot be told apart. Metformin use was not reliably ascertainable in our intake data, so we did not model it; it is part of the residual confounding rather than a variable we controlled. The largest *measurable* selection variable, insurance, was included directly in the matching and was well balanced afterward (Table 1). True socioeconomic status is unobserved, so residual confounding correlated with both formulation choice and outcome remains possible. The equivalence result is, however, unlikely to be an artifact of unmeasured confounding: the unadjusted difference is already small, and matching on the measured covariates (insurance included) leaves the formulations equivalent within the pre-specified margin.

## 4. Discussion

### 4.1 What this paper says, plainly

Branded and compounded tirzepatide were statistically equivalent in six-month effectiveness within a pre-specified ± 2 pp margin (TOST *p* < 0.001) in this telehealth cohort. The unadjusted edge of branded was small to begin with (+0.20 pp on loss; +2.1 pp on response) and reflects who chooses branded, chiefly the more-insured patients (the one large baseline imbalance), rather than the formulation; matching on insurance and the other baseline covariates leaves the two equivalent.

The cost advantage of compounded is real but conditional on the branded price the patient actually pays. The list-price comparison that the cost literature has typically anchored to overstates the savings by roughly fourfold relative to LillyDirect Self Pay, and the savings turn negative, with compounded costing more, for patients with insurance copays below $200/month. Because effectiveness is equivalent, the decision reduces to price: compounded is cheaper for patients without good branded access, but it is not a default lower-cost substitute across all coverage tiers.

### 4.2 What we cannot conclude

We demonstrated statistical equivalence in six-month *weight-loss outcomes* within a ±2 pp margin; this is not the same as pharmacologic bioequivalence. The equivalence holds under the identifying assumptions encoded in the DAG, and some of those (metformin not lumping confounder and mediator, no unobserved confounders correlated with both formulation choice and outcome) are not testable with our data. A formal bioequivalence study with a different design is the appropriate evidence for the pharmacologic question. The ± 2 pp margin is a clinical judgment; a reader who regards a smaller difference as meaningful should interpret the result against that margin.

We cannot conclude that all compounded preparations are equivalent. We dropped the formulation-additive subanalysis because we could not separate “additive effect” from “compounding-pharmacy fingerprint” in our data; a by-pharmacy analysis with explicit pharmacy identifiers is the appropriate way to address it.

We cannot conclude anything about adverse-event rates. Our outcome is weight loss; safety is a separate question requiring the appropriate exposure denominators and adverse-event ascertainment.

### 4.3 Why the price-tier reframing matters

Cost analyses for compounded GLP-1 therapy that anchor on retail list price implicitly assume the comparator is a patient with no insurance and no manufacturer-direct access. In 2026 that assumption is increasingly false: LillyDirect Self Pay is widely available to cash patients, and a substantially higher share of our branded cohort reported insurance coverage than the compounded cohort (the dominant baseline difference between the groups). The literature should report price-conditional cost comparisons, not single-anchor savings claims.

### 4.4 Limitations

#### Selection bias

The unadjusted gap demonstrates that selection is real. PSM addresses selection on observed covariates only. The DAG identifies metformin and SES as residual concerns, which we address via stratification and sensitivity but cannot fully eliminate.

#### No bioequivalence claim

See Section 4.2.

#### Single-platform retrospective

The cohort is from one telehealth provider’s data; external validation in other compounded-GLP-1 programs is needed before generalizing the finding of no detected effectiveness difference.

#### Cost comparison, not cost-effectiveness analysis

We report dollar cost differences only. We computed no incremental cost-effectiveness ratio (ICER), no quality-adjusted life years (QALYs), and no discounting; the analysis is a cost comparison (cost-minimization under the assumption of similar effectiveness), not a formal cost-effectiveness analysis.

#### Price scenarios are point estimates

LillyDirect Self-Pay Journey prices ($299/$399/$449) are current as of the February 2026 program and may change. Insurance copays vary widely by plan; we use illustrative ranges rather than comprehensive distributions. Compounded savings are computed against a single $200/month anchor and would shrink against a higher compounded price such as the cited market-median (approximately $300/month).

#### Outcome ascertainment

Patients with a 6-month observation may be more engaged than the full prescribed cohort. We restrict to patients with documented 6-month weights in both arms; differential ascertainment between arms is not formally addressed.

## 5. Conclusions

In a real-world telehealth cohort of tirzepatide users (869 branded-only and 6,402 compounded-only patients with a six-month weight, 718 matched pairs; switchers excluded), branded and compounded tirzepatide were statistically equivalent in six-month weight-loss outcomes within a pre-specified ± 2 pp margin (TOST *p* < 0.001), after matching on baseline covariates including insurance. Because effectiveness is equivalent, the choice is a cost decision, and the cost advantage of compounded is real but conditional on the branded price tier the patient can access: $6,000 in six-month savings versus retail list price, approximately $594–$1,494 versus LillyDirect Self Pay (2.5 mg and maintenance-dose tiers respectively), and approximately zero or negative for patients with insurance copays below $200/month. The cost literature should adopt price-conditional reporting rather than anchoring on a single retail-list comparator.

## Data Availability

Individual patient-level data are private and cannot be shared publicly. De-identified data and the analysis code can be provided upon reasonable request to the corresponding author.

## Author Contributions

**Brian Erly:** Conceptualization, DAG development, Supervision, Writing, Clinical validation. **Shanmugesh Raja:** Conceptualization, Data curation, Formal analysis, Methodology, Software, Visualization, Writing.

## Ethics Approval

This study is a secondary analysis of de-identified data collected during routine clinical care at Mochi Health. As the analysis involves no direct patient contact, no intervention, and uses only data de-identified per HIPAA Safe Harbor [45 CFR 164.514(b)(2)], it was determined to not constitute human subjects research as defined by 45 CFR 46.102. Formal IRB review was therefore not required.

